# A recurrent neural network and parallel hidden Markov model algorithm to segment and detect heart murmurs in phonocardiograms

**DOI:** 10.1101/2023.12.26.23300540

**Authors:** Andrew McDonald, Mark JF Gales, Anurag Agarwal

## Abstract

Detection of heart disease using a stethoscope requires significant skill and time, making it expensive and impractical for widespread screening in low-resource environments. Machine learning analysis of heart sound recordings can improve upon the accessibility and accuracy of diagnoses, but existing approaches require further validation on larger and more representative clinical datasets. For many previous algorithms, segmenting the signal into its individual sound components is a key first step. However, segmentation algorithms often struggle to find S1 or S2 sounds in the presence of strong murmurs or noise that significantly alter or mask the expected sound. Segmentation errors then propagate to the subsequent disease classifier steps. We propose a novel recurrent neural network and hidden semi-Markov model (HSMM) algorithm that can both segment the signal and detect a heart murmur, removing the need for a two-stage algorithm. This algorithm formed the ‘CUED Acoustics’ entry to the 2022 George B. Moody PhysioNet challenge, where it won the first prize in both the challenge tasks. The algorithm’s performance exceeded that of many end-to-end deep learning approaches that struggled to generalise to new test data. As our approach both segments the heart sound and detects a murmur, it can provide interpretable predictions for a clinician. The model also estimates the signal quality of the recording, which may be useful for a screening environment where non-experts are using a stethoscope. These properties make the algorithm a promising tool for screening of abnormal heart murmurs.

**Author summary:** The use of machine learning algorithms to detect heart disease from sound recordings has great potential to enable widespread and low-skill screening, improving early detection and treatment. The area has seen increasing interest in recent years, with many novel algorithms inspired by deep learning advancements in other fields. However, the size of heart sound datasets remains small, making deep learning models particularly susceptible to overfitting. In addition, the performance of these algorithms has rarely been directly compared on unseen data. We describe a novel lightweight algorithm to detect and classify murmurs in heart sound recordings. This algorithm was the winning entry into the George B. Moody PhysioNet 2022 challenge, beating many complex deep-learning approaches. Our approach both detects and localises the murmur, providing an interpretable result for a clinician.

## Introduction

Cardiovascular disease is the leading cause of mortality worldwide, with 18 million deaths every year [1]. However, compared to other serious diseases, awareness of many types of heart disease remains low and many are critically underdiagnosed [2]. In young people, rheumatic valvular heart disease is the most common cardiovascular disease, with an estimated global burden of 41 million cases, primarily in developing countries [3]. More than a quarter of patients with rheumatic disease present to a clinician at a late stage with heart failure [4]. There is a clear need for widespread early detection, to improve early treatment of the disease and prevent long-lasting morbidity [1].

Many heart diseases, such as rheumatic valve disease, cause structural changes in the heart that lead to abnormal sounds, such as heart murmurs [5]. The only tool currently available to detect these sounds in primary care is a stethoscope. Listening to the chest with a stethoscope (auscultation) is a quick and non-invasive test. However, auscultation proficiency varies widely amongst clinicians. The sensitivity of an experienced general practitioner in detecting valvular heart disease can be as low as 44% [6]. Enabling low-cost screening of the disease, especially in resource-constrained areas, will require a test that can be quickly and accurately performed by non-specialist clinicians.

Automated analysis of heart sound recordings (phonocardiograms) is a promising solution to improve the accuracy and accessibility of auscultation. A number of novel methods have been proposed in recent years, driven by an increased amount of open-access datasets and renewed interest in machine learning and AI [7]. However, many of these algorithms suffer from issues such as overfitting and poor generalisation to new data. The George B. Moody PhysioNet 2022 challenge [8] tasked participants to design novel algorithms to detect and classify heart murmurs in a new paediatric dataset, enabling an independent comparison of approaches that is representative of a real-world clinical environment.

In this paper, we describe a novel algorithm that won the First Prize in both tasks in the challenge [9, 10]. The algorithm is inspired by takeaways from the earlier 2016 PhysioNet challenge [7, 11] and, in contrast to many other approaches, does not use an end-to-end deep learning model. We also explore the results of the challenge and compare the algorithm’s efficacy on the two distinct tasks.

## Materials and methods

### Availability of data

An ongoing limitation of research into automated analysis of heart sound recordings (also known as phonocardiograms, PCGs) is the availability of high-quality labelled data. Whilst electronic stethoscopes have received regulatory approval and are available to purchase, the vast majority of stethoscopes in clinical practice are analogue. Therefore, heart sounds are rarely recorded and stored with patient records, unlike other cardiac tools such as electrocardiography (ECG) and echocardiography. The creation of heart sound datasets therefore requires bespoke clinical research studies that are expensive and time-consuming, especially in resource-strained cardiology units.

Due to these constraints, open-access data has proved a valuable resource for advancing algorithm design. These datasets include the 2016 PhysioNet/Computing in Cardiology dataset [12] and the PASCAL Challenge dataset [13]. However, these datasets are limited by a lack of detailed information on the recording environment, murmur assessments, and patient outcomes. The 2016 PhysioNet challenge included a withheld test set to assess the generalisation of algorithms [7]. However, this test set was never made public after the challenge. Research using this dataset since 2016 has created new test sets by applying varying cross-validation and splitting strategies to the training set. These varied strategies have made direct comparisons between approaches challenging. The 2016 PhysioNet dataset also used recordings from multiple different devices, with its largest dataset using different devices to record abnormal and normal patients [12]. This unintentionally prompted machine learning algorithms to overfit to the dataset by learning to distinguish device characteristics rather than diagnostic sounds [11].

### The George B. Moody PhysioNet Challenge 2022

To address the challenges of data availability and algorithm comparison, The George B. Moody PhysioNet Challenge 2022 [8] tasked participants to design algorithms to detect heart murmurs and predict clinical outcomes in the new CirCOR paediatric dataset [14].

As described by Oliveira et al. [14], the dataset was gathered as a part of two screening programs in Brazil in 2014 and 2015. Approval for the study protocol was granted by the 5192-Complexo Hospitalar HUOC/PROCAPE Institutional Review Board under the request of the Real Hospital Português de Beneficiência em Pernambuco. Written consent was obtained for all participants, with parental consent where appropriate.

A total of 5268 phonocardiogram recordings were collected from 1452 patients. Some patients were recruited more than once, giving 1568 unique patient encounters. Recordings were made using the Littmann 3200 electronic stethoscope at up to four of the standard auscultation sites on the chest (aortic, pulmonic, tricuspid, mitral).

For the challenge, 60% of the data was released publicly, with the remaining 40% split between a validation (10%) and a final test set (30%) [10]. Participants were able to submit their algorithms to the validation set throughout the challenge to assess their performance, whilst the challenge organisers ran each entry only once on the test set to determine the final ranking [10].

In this work, we train and analyse algorithms using the PhysioNet 2022 dataset, reporting results on both the training and withheld test sets. This dataset is publicly available at the Challenge website [15].

### Previous Work

Whilst the PhysioNet 2022 dataset is a significant addition to the size of open-access PCG data, it is still small compared to many machine learning domains where deep learning algorithms dominate performance. Feature extraction is a beneficial step to reduce the complexity of the audio data and hence the complexity required of subsequent classifiers, making it easier to train them to identify diagnostic features. A key conclusion of the 2016 PhysioNet challenge was that feature extraction was the most ‘crucial and important’ part of algorithms [7].

The nature of heart sounds is well-understood from a clinical perspective [5]. A phonocardiogram is a non-stationary signal consisting of a generally periodic set of S1 (lub) and S2 (dub) sounds corresponding to closures of the atrioventricular and semilunar valves respectively [5]. Abnormal murmurs can appear in the systolic and diastolic regions of the signal, depending on the particular valve pathology of the patient. Other abnormal sounds, such as S3 and S4, may also appear, giving distinctive rhythms in the signal. Due to this periodic structure, a common first step in classifiers is segmentation, where the individual heart sound states (S1, S2, systole, diastole) are labelled in time. This allows subsequent algorithm stages to focus on diagnostically relevant areas of the recording and apply ensemble averaging to segments to reduce noise.

In the 2016 challenge, a segmentation algorithm designed by Springer et al. [16] was provided to participants to aid their design [7]. This model used a hybrid structure, where a logistic regression provided observations for a hidden semi-Markov model (HSMM). It was considered state-of-the-art at the time [7]. However, the algorithm assumes a healthy heart cycle which makes it susceptible to errors when loud murmurs or noise overwhelm weaker S1 or S2 sounds [17].

Kay [17] observed this limitation and designed a segmentation algorithm that directly models the heart sound state. He calculates a series of band-pass-filtered homomorphic envelopes and power spectral densities (PSDs) to give a feature set that can distinguish murmurs from healthy sounds. He then replaces the logistic regression of Springer with a fully connected neural network, giving greater non-linear discrimination between states. Noting that the algorithm of Springer struggles to distinguish S1 from S2 and systole from diastole, Kay’s algorithm predicts only three states: murmur, major heart sound, and silence. These neural network predictions are used as observations for two hidden semi-Markov models, one of which assumes the murmur state appears in systole and one that assumes a healthy heart sound with silence in systole. The most confident segmentation produced by the HSMM gives a murmur diagnosis. We provide more detail on this HSMM approach in the Methods section of this paper.

One limitation of the approaches of Springer [16] and Kay [17] is that the discriminative classifiers providing observation probabilities predict on a per-frame basis and do not model dependencies between adjacent time frames. When inspecting a phonocardiogram, a skilled clinician will identify the S1 and S2 sounds by assessing the timing of the sounds. The models of Springer and Kay cannot do this as they view each frame independently, and so rely on the HSMM to provide this timing information. In this work, we deploy a recurrent neural network (RNN) to model this time dependency. RNNs have been applied for heart sound segmentation [18], offering improved predictive power compared to logistic regressions. However, the models deployed in previous work neglect to predict a murmur state and assume that only a healthy heart sound cycle is present.

### Method

The algorithm we describe in the following sections is an improvement over Kay’s work [17] and has been optimised for the 2022 PhysioNet challenge. The algorithm, as shown in Fig 1, consists of four distinct stages: (i) feature extraction, (ii) neural network prediction, (iii) segmentation, and (iv) final classification through a comparison of segmentation confidences.

**Fig 1.**
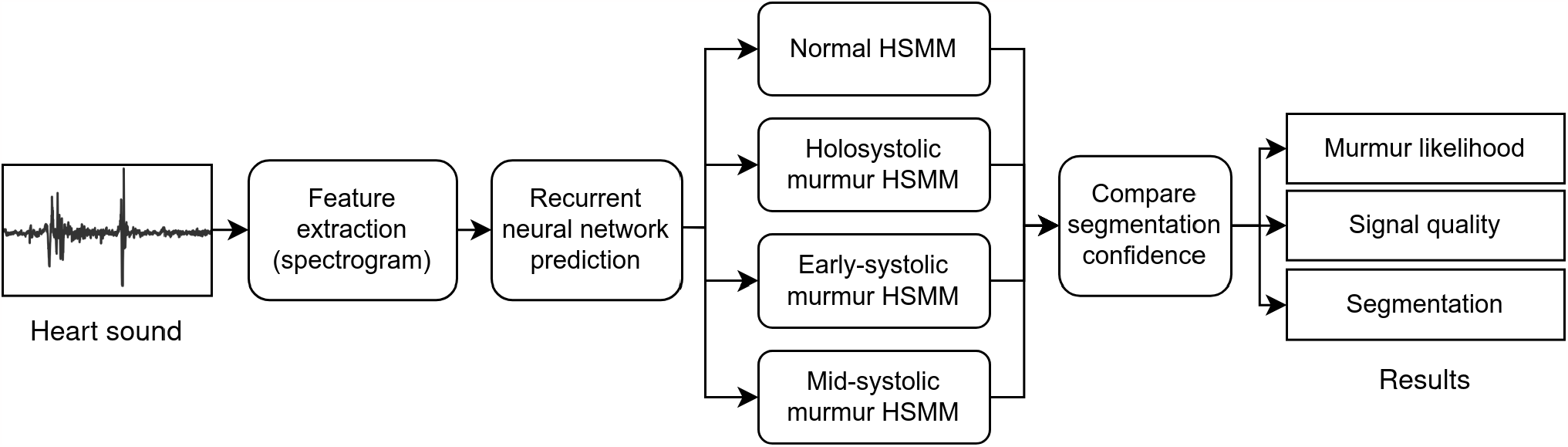
Parallel HSMM algorithm structure. A heart sound recording is transformed into log-spectrogram features, which are then input to an RNN. The RNN probabilities are then used to produce four segmentations using four parallel HSMMs. The confidence in these segmentations is then compared to give the most probable segmentation, murmur prediction, and a signal quality estimate.

We define a time-series heart sound recording with *N* samples as *r*_1:*N*_. All the recordings in the challenge dataset were made using the Littmann 3200 electronic stethoscope, which has a fixed sampling frequency of 4000 Hz. In feature extraction, the time-series recording is converted into a time-frequency series with *T* time windows, *x*_1:*T*_. Four parallel HSMM models (*ω*_1_, …, *ω*_4_), are then applied to produce four distinct segmentations 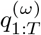. The confidence in these segmentations is then compared to produce a final segmentation and classification.

The following subsections describe each step of the algorithm in more detail. All computation was conducted in Python 3 using PyTorch 1.11, NumPy 1.21, and SciPy 1.7. The code to train these models and generate results is available on a public repository [19]. The corresponding datasets are available at the PhysioNet 2022 challenge website [15, 20].

### Feature Extraction

The amplitude of the PCG is first normalised by removing its mean and dividing by the resulting peak amplitude. The absolute amplitude of a PCG is an unreliable feature as it varies significantly depending on the application pressure of the stethoscope and the particular patient [21].

The homomorphic envelope and PSD features of Kay [17] allow discrimination of energy in high and low frequencies but there is significant redundancy and extra computation in computing both the band-pass-filtered envelopes and the PSDs. Instead, in this work we compute a log-spectrogram using a Hann window of length 50 ms and step 20 ms. A larger window length enables a higher frequency resolution at the cost of a lower time resolution. The time duration of S1 and S2 sounds is approximately 100 ms, so a 50 ms window was chosen to enable precise identification of the major heart sounds whilst maintaining an effective frequency resolution of 20 Hz. A secondary advantage of this approach is that spectrograms provide an interpretable 2D visualisation of the time-frequency energy in the recording, as shown in Fig 2.

**Fig 2.**
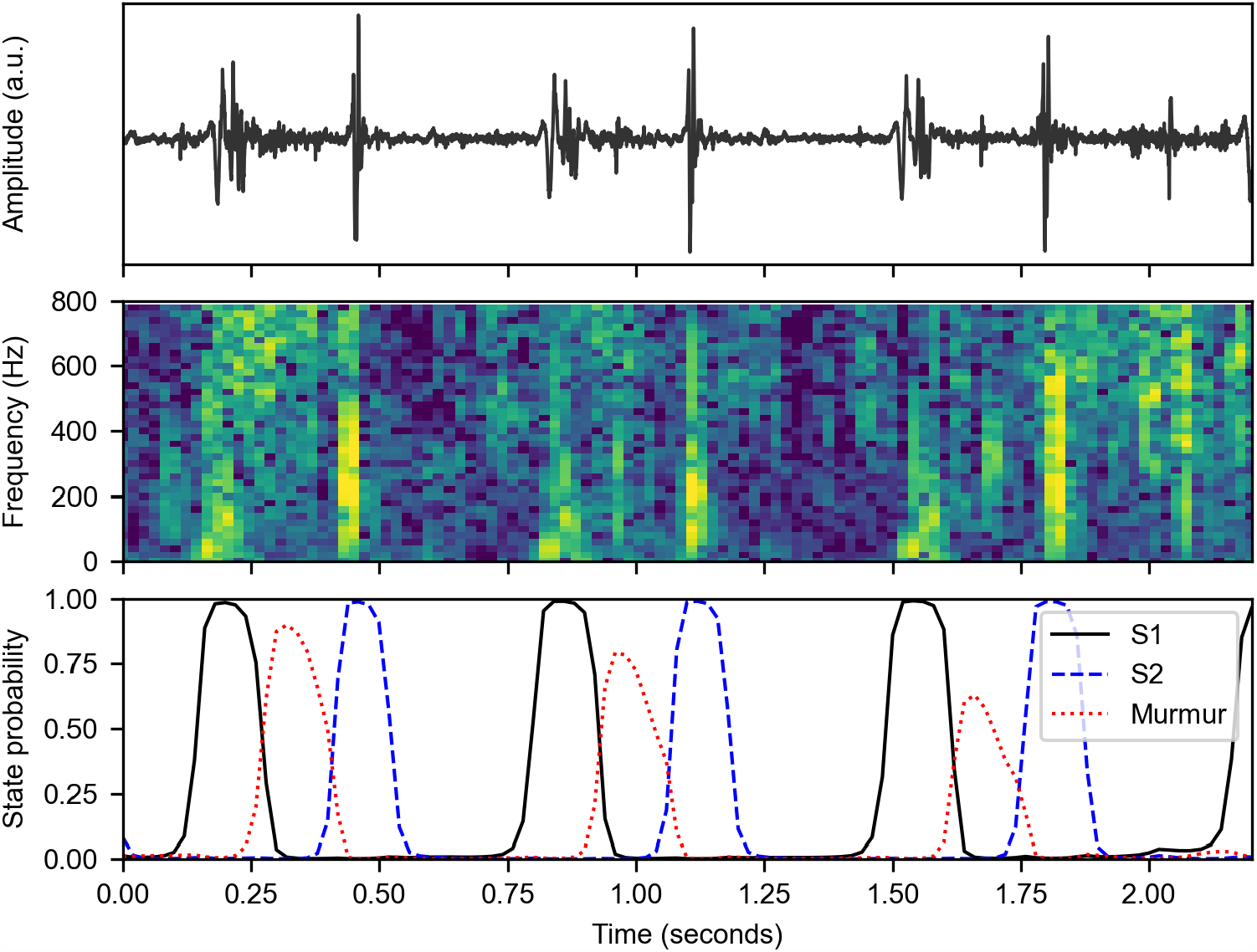
Neural network prediction of per-time heart sound categories. The heart sound recording (top) is transformed to a normalised spectrogram (middle) that is then passed to a RNN that predicts the state corresponding to each time window (bottom). The RNN is able to use the spectrogram to correctly distinguish between S1, S2, and the systolic murmur. The RNN also predicts systolic and diastolic states which are omitted from the bottom figure for clarity.

Although the Nyquist frequency of the recordings is 2000 Hz, we further limit the spectrogram to 0-800 Hz to remove higher frequencies that contain no heart sound information. This reduces the risk of the subsequent neural network stages learning to overfit to irrelevant high-frequency noise such as speech and background sounds.

As a key final step, each frequency row in the spectrogram is z-score normalised by subtracting its mean and dividing by its variance. Murmurs commonly contain much less time-frequency energy than S1 and S2 sounds, and this normalisation reduces the dynamic range [11].

### Recurrent Neural Network

A recurrent neural network with knowledge of the timing of heart sounds should be able to discriminate S1, S2 and murmur sounds without relying on a subsequent HSMM. We therefore define a ground-truth segmentation *q*_1:*T*_ of five distinct heart sound states *ξ*_*i*_ ∈ {S1, S2, systole, diastole, murmur}. The challenge dataset includes segmented locations for the S1, S2, systole and diastole sounds but does not explicitly localise murmurs. However, the additional labels provided by the challenge include a prediction of the ‘murmur timing’ annotated by a clinician [14]. We use these labels to approximately annotate the location of the murmur in the ground-truth segmentation.

If the recording is labelled to contain an early-systolic murmur, the first 50% of each systolic period is annotated as ‘murmur’. If ‘mid-systolic’, the middle 50% is murmur. If ‘holosystolic’, the entire systole portion is annotated as a murmur.

We note that one limitation of this analysis is that diastolic murmurs are not considered. Diastolic murmurs are much less prevalent than systolic murmurs in clinical practice, and only 5 patients in the dataset have diastolic sounds. A future improvement, given more diastolic examples, would be to replicate this labelling approach for diastolic murmur signals.

Using this modified ground-truth segmentation, we train a bidirectional RNN with parameters *θ* to predict the state *q*_*t*_ at each time instance using the log-spectrogram features *x*_1:*T*_, giving posterior probabilities *P* (*q*_*t*_ = *ξ*_*i*_| *x*_1:*T*_, *θ*). An example of the outputs of the RNN is shown in Fig 2. The RNN is confidently able to distinguish S1 and S2, verifying the five-state segmentation model.

The RNN structure and hyperparameters are optimised through cross-validation on the training dataset. The final model consists of a three-layer bidirectional RNN with Gated Recurrent Unit (GRU) [22] cells with a hidden layer size of 60. The concatenated forward and backward outputs of the RNN are passed to a two-layer fully connected neural network with Tanh activations and hidden sizes of 60 and 40. This fully connected network and a subsequent softmax layer reduce the RNN output to the five-dimensional output of the segmentation labels. Dropout with probability 0.1 is applied between the GRU and fully connected layers to reduce overfitting.

The model is trained using a cross-entropy loss with the Adam optimiser [23]. Some states in the segmentation (e.g. diastole) are much more prevalent than others (e.g. murmur), so to avoid models learning to favour one class the loss function is inversely weighted by the frequency of each class label in the overall dataset.

### Parallel Hidden Semi-Markov Models

Given posterior probabilities from the RNN, the simplest method to produce a segmentation would be to ‘greedily’ pick the state with the maximum probability at each time instance:

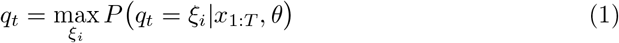

A murmur could then be predicted if the greedy segmentation ever contains a murmur state. However, in practice, this approach is very prone to false positive murmur predictions because higher-frequency signal noise can cause occasional spurious predictions. Additionally, the heart is physiologically constrained to generate sounds in the order S1, systole, S2, diastole but the RNN is not similarly bound. This means that physically impossible state transitions are possible (e.g. S2 to systole) in the greedy segmentation.

To generate a globally optimum and physically valid segmentation we use the RNN probabilities as observations for hidden semi-Markov models, following the hybrid structure used in Springer [16] and Kay [17].

As described by Springer et al. [16], the HSMM is an extension to a traditional hidden Markov model that uses an explicit model for the duration of each state. This is particularly useful for physiological signals, such as phonocardiograms, where states have reasonably well-defined durations due to physical constraints.

The expected durations of states (particularly systole and diastole) significantly vary between patients due to the wide range of resting heart rates in the dataset. Springer et al. [16] therefore does not use one global model for the state durations, instead fine-tuning Gaussian distributions by scaling their means by an estimate of the heart rate. Their approach follows the work of Schmidt et al. [24] and estimates the heart rate by computing the autocorrelation of a smoothed homomorphic envelope of the heart sound signal. They then search for the highest peak in the autocorrelation in the 500 to 2000 ms range, corresponding to heart rates between 30 bpm and 120 bpm. This search range requires modification for paediatric use as approximately 20% of the dataset has a heart rate above 120 bpm.

In this work, we improve upon this estimate of the heart rate by noting that the posterior probabilities from the RNN are a filtered version of the original signal from which a period can be estimated. The homomorphic envelope Springer uses can therefore be replaced with the RNN posteriors to get an autocorrelation that is much smoother and less affected by noise spikes. See S1 Fig for more information.

Kay uses two HSMMs, one assuming a healthy sound and the other assuming a holosystolic murmur [17]. However, many of the murmurs in this dataset are early systolic [14]. In this work, we improve upon these assumptions by using four HSMMs that each make different assumptions about the underlying signal and hence generate a different segmentation. The state durations are shared between each HSMM but the observation probabilities and transition matrix differ:

*ω*_1_ A normal signal with no murmur. A four-state segmentation model is used with the murmur posterior from the RNN discarded.

*ω*_2_ A holosystolic murmur signal. A four-state segmentation model is used, where the murmur posterior replaces every systole posterior.

*ω*_3_ An early-systolic murmur signal. A five-state segmentation model is used, with a transition matrix that forces the S1 state to transition to the murmur state and then to the systolic state.

*ω*_4_ A mid-systolic murmur signal. A five-state segmentation model as above, but with a transition matrix that forces the S1 state to go to systole first.

A predicted segmentation 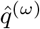 is calculated using each HSMM model above, giving four distinct interpretations of the signal. We then calculate a confidence measure of the segmentation *C*^(*ω*)^ by tracing the predicted segmentation path back through the RNN posterior probabilities:

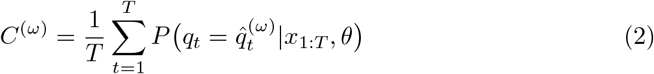

The final model, 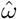, is chosen as the HSMM with the largest confidence:

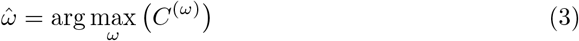

This selects a final predicted segmentation of the signal, 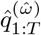, and a final classification of the type and location of any systolic murmur. The confidence of the chosen model, 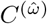, can be used as a measure of the signal quality. The four-class model can easily be reduced to a binary murmur detector; if the chosen model is a murmur HSMM model 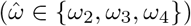 then a murmur is predicted. For the binary case, we can also calculate an overall confidence of the murmur, *C*^(*M*)^, vs no murmur, *C*^(*N*)^ decision:

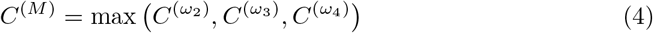

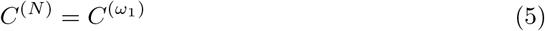

The difference between these two confidences can then be used as a measure of the separability of the murmur and no murmur outcomes:

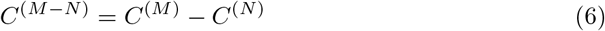

The above method produces a per-recording prediction of the presence of a murmur. To get a per-patient prediction in the format required by the challenge, we apply a simple common-sense criterion. If a murmur is detected in any of the recordings, the patient is predicted to have a murmur. If this is not true, the confidence of the chosen model 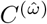 is examined: if this is below a threshold (0.65), the patient is classified as ‘unknown’. Otherwise, the patient is predicted to be ‘no murmur’.

### Prediction of Clinical Outcome

The second task of the challenge was to predict the final clinical outcome of the patient. A skilled clinician will auscultate each location of the chest and weigh the strength and characteristics of the sounds at each site when trying to predict clinical significance [5]. They may also consider general patient biometrics such as age, and sex, along with the recorded patient history.

The dataset provided in the challenge does not include detailed patient biomarkers or risk factors but does include basic biometrics such as age. To incorporate this information alongside the heart sound recordings, we apply a CatBoost gradient boosted decision tree [25] as shown in Fig 3. For each recording, we use the parallel HSMM method above to calculate the murmur likelihood, *C*^(*M*−*N*)^. We also compute the maximum confidence, 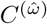, so that the decision tree can reject poor-quality signals.

**Fig 3.**
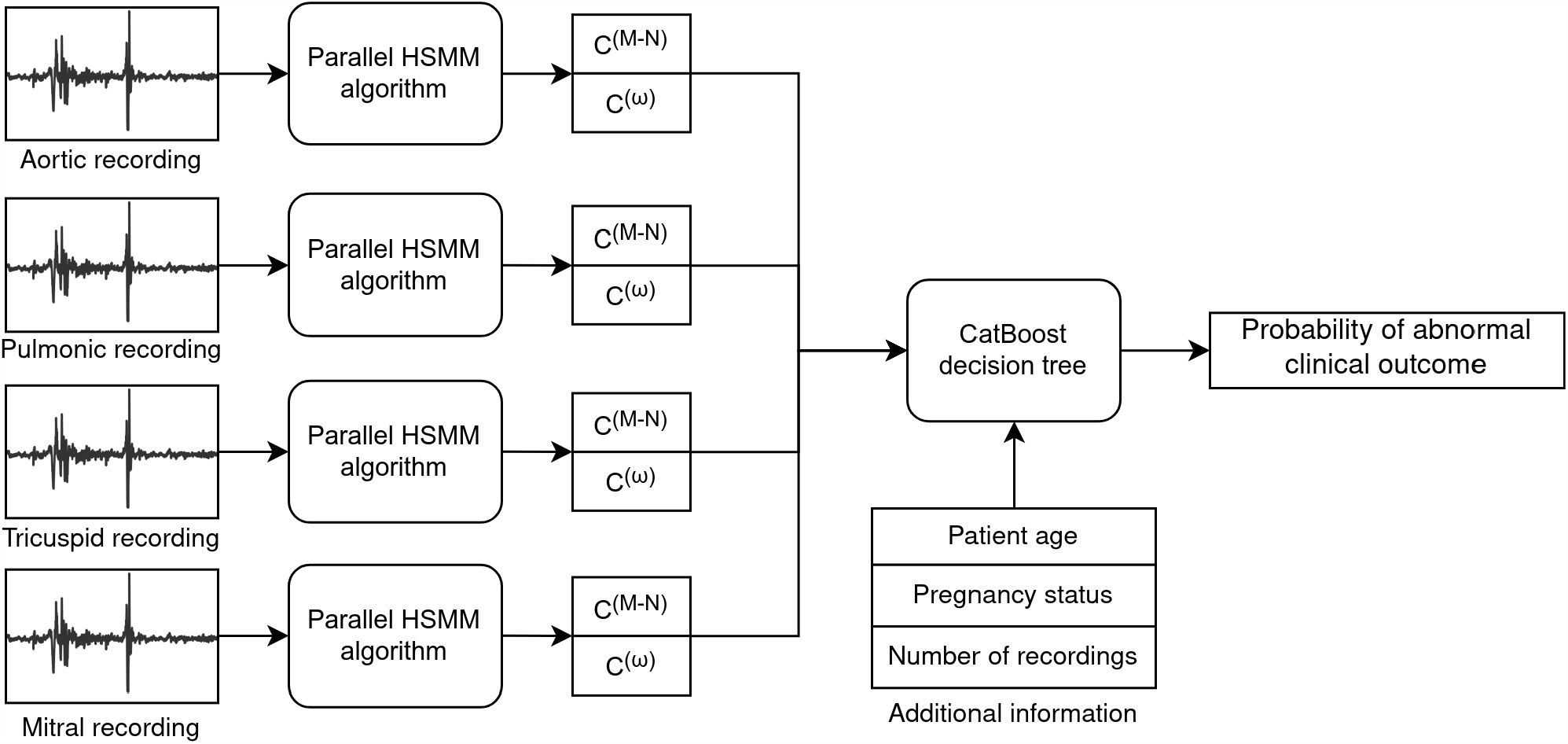
CatBoost algorithm structure to predict clinical outcome. The parallel HSMM algorithm described in Fig 1 is applied to each recording to generate murmur likelihood *C*^(*M*−*N*)^ and signal quality 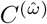 metrics. These metrics are then combined with additional patient information and input to a CatBoost decision tree to predict the patient’s final clinical outcome.

In situations where a patient has multiple recordings from the same chest location, the values are averaged before input to the CatBoost model. Added to the feature set are the patient’s age, pregnancy status, and the overall number of recordings made.

The CatBoost model is trained with a cross-entropy loss and optimised through a five-fold cross-validation strategy alongside the RNN model. A class weight of 1.8 is used for the abnormal examples and 1 for the normal examples because the challenge cost function prioritises sensitivity. The chosen decision tree has a depth of 9. The final threshold probability to decide an abnormal result (0.4738) is chosen to minimise the challenge cost function.

## Results and Discussion

### Official PhysioNet challenge results

Table 1 provides the official results of our algorithm on both challenge tasks [10]. A total of 40 teams competed in the competition and received official scores. The algorithm achieved the second-highest score in the murmur detection task, with an accuracy just 0.004 below the top score. In the clinical outcomes task, the algorithm achieved the top score. Some teams’ entries failed subsequent tests to their code which made them ineligible for final prizes [10]. The described algorithm therefore won the First Prize in both tasks.

**Table 1.** Official challenge results [15] for parallel HSMM algorithm.

| Task | Training | Validation | Test | Final Rank |
| --- | --- | --- | --- | --- |
| Murmur detection accuracy | 0.817 | 0.758 | 0.776 | 2/40 |
| Clinical outcome score | 10565 | 9257 | 11144 | 1/40 |

The ranking of entries changed significantly between the validation and test sets. The final scores (Table 1) also showed a significant difference between the validation and other sets. The validation set (10% of the data) was smaller than the training set (30%) and teams were allowed up to 10 submissions to the validation set. Teams, therefore, optimised their algorithm for best performance on the validation set, which, given the relatively small total dataset, was not entirely representative of the final test set. In particular, the prevalence of the ‘abnormal outcome’ in the validation set (0.383) was significantly lower than in the test set (0.507) [10].

The metric for murmur detection is an accuracy that is weighted to penalise false negatives. The score for the clinical outcomes task is a custom loss function for the challenge, where lower scores are better.

## Cross-validated results

The test set was not made available to teams after the challenge. This means only a limited set of result metrics can be computed. For an in-depth exploration of the algorithm, we additionally report additional metrics evaluated on the public training set. These results are evaluated via a 5-fold cross-validation procedure.

### Murmur detection

Fig 4 shows a plot of the HSMM confidence values for every recording in the training dataset. Using the HSMM confidences allows for a strong separation of murmur and normal signals, whilst producing an estimate of signal quality. Fig 5 shows a reliability diagram for murmur detection, where the murmur likelihood (*C*^(*M*−*N*)^) is plotted against the relative frequency of murmurs. The approximately directly proportional relationship shows that the murmur likelihood provides a calibrated estimate of the confidence in the decision. A low threshold of *C*^(*M*−*N*)^ = 0 was chosen for the challenge because of the weighted accuracy penalising false negatives. However, a higher threshold could be picked for future applications, such as population screening, where a high specificity is essential to minimise false positive referrals.

**Fig 4.**
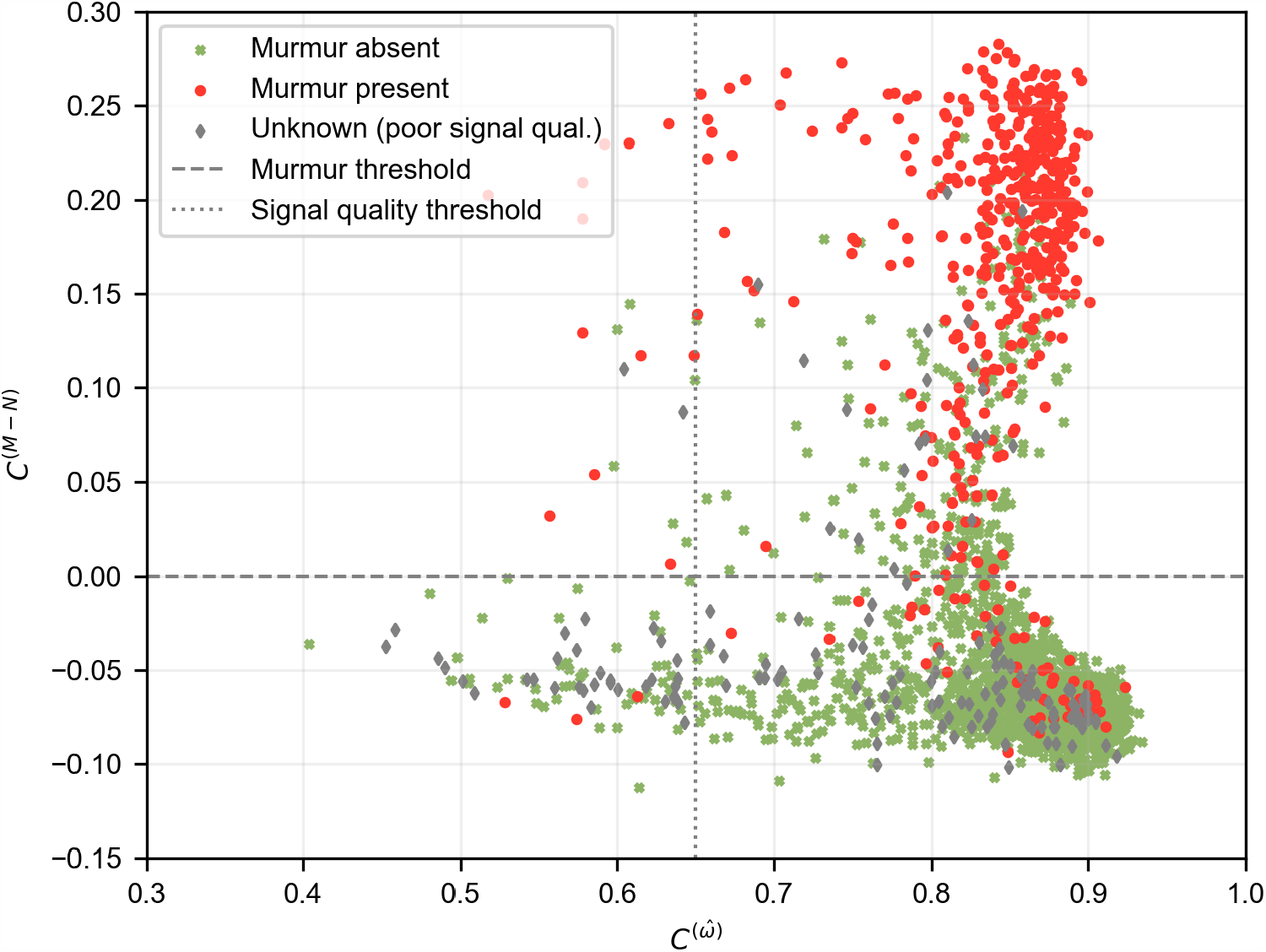
Separation of murmur, normal, and poor signal quality signals using HSMM confidences. The difference between the normal HSMM confidence, *C*^(*N*)^, and the most confident murmur HSMM, *C*^(*M*)^, is used to predict murmur likelihood *C*^(*M*−*N*)^, whilst the most confident overall HSMM 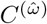 is used as an indication of signal quality. The horizontal and vertical lines show thresholds that have been chosen to separate the different classes, optimised for the challenge task. The marker type indicates the ground-truth labels.

**Fig 5.**
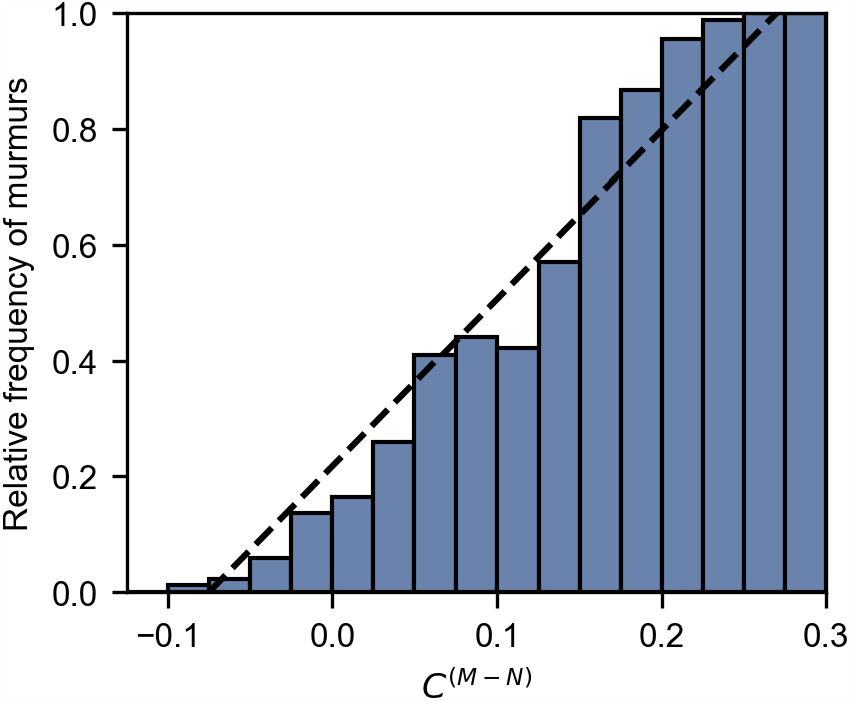
Reliability diagram for prediction of murmurs using HSMM confidence difference. As the confidence difference *C*^(*M*−*N*)^ increases, so does the relative frequency of murmurs with an almost directly proportional relationship (dashed line). This suggests good calibration of the murmur likelihood output.

One limitation of this dataset is that the murmur labels were annotated by a single clinician rather than a board of reviewers [10]. The labelling of a heart murmur depends heavily on the annotator’s skill, hearing acuity, and headphones. Therefore, it is not as repeatable a ground-truth as other cardiac tests such as electrocardiography and echocardiogram and some disagreements between the algorithm and the clinician label are to be expected. Future work should investigate recordings where a large majority of challenge teams disagree with the clinician label, as this may be indicative of mislabels.

Table 2 shows the per-class performance of the algorithm in terms of sensitivity and positive predictive value (PPV). The algorithm at its current operating point is 92.7% sensitive to murmurs, which leads to a high challenge weighted accuracy because of the heavy penalty applied to false negatives. As expected, the sensitivity of the algorithm increases with the patient’s reported murmur grade. For grade 1 (quiet) murmurs, 87.5% of cases are detected. This rises to 100% for grades 2 (moderate) and 3 (loud). The algorithm also has a high precision for the ‘murmur absent’ class, which would be important for use as a rule-out device where patients must be confidently rejected as having a murmur. The performance at predicting the ‘unknown’ class is poor. This class was used if the annotator was unable to confidently predict the presence of a murmur [14]. However, this extra label is subjective and highly dependent on how the recordings were listened to and annotated. The definition of a poor-quality signal from an algorithmic and human perspective is likely very different. It is possible that the algorithm can confidently predict cases that a human cannot, due to analysis of lower-frequency inaudible energy or greater resilience to noise. Fig 6d shows an example of one of these recordings, which does contain some noise and is marked unknown by the clinician, but still has audible heart sounds and is confidently segmented by the algorithm and predicted as ‘Murmur absent’.

**Table 2.** Per-class results for the murmur detection task.

| Class | Cases | Sensitivity (%) | PPV (%) | $F_1$ score |
| --- | --- | --- | --- | --- |
| Murmur present | 179 | 92.7 | 55.0 | 0.692 |
| Unknown | 68 | 30.9 | 34.4 | 0.328 |
| Murmur absent | 695 | 77.6 | 93.1 | 0.848 |

**Fig 6.**
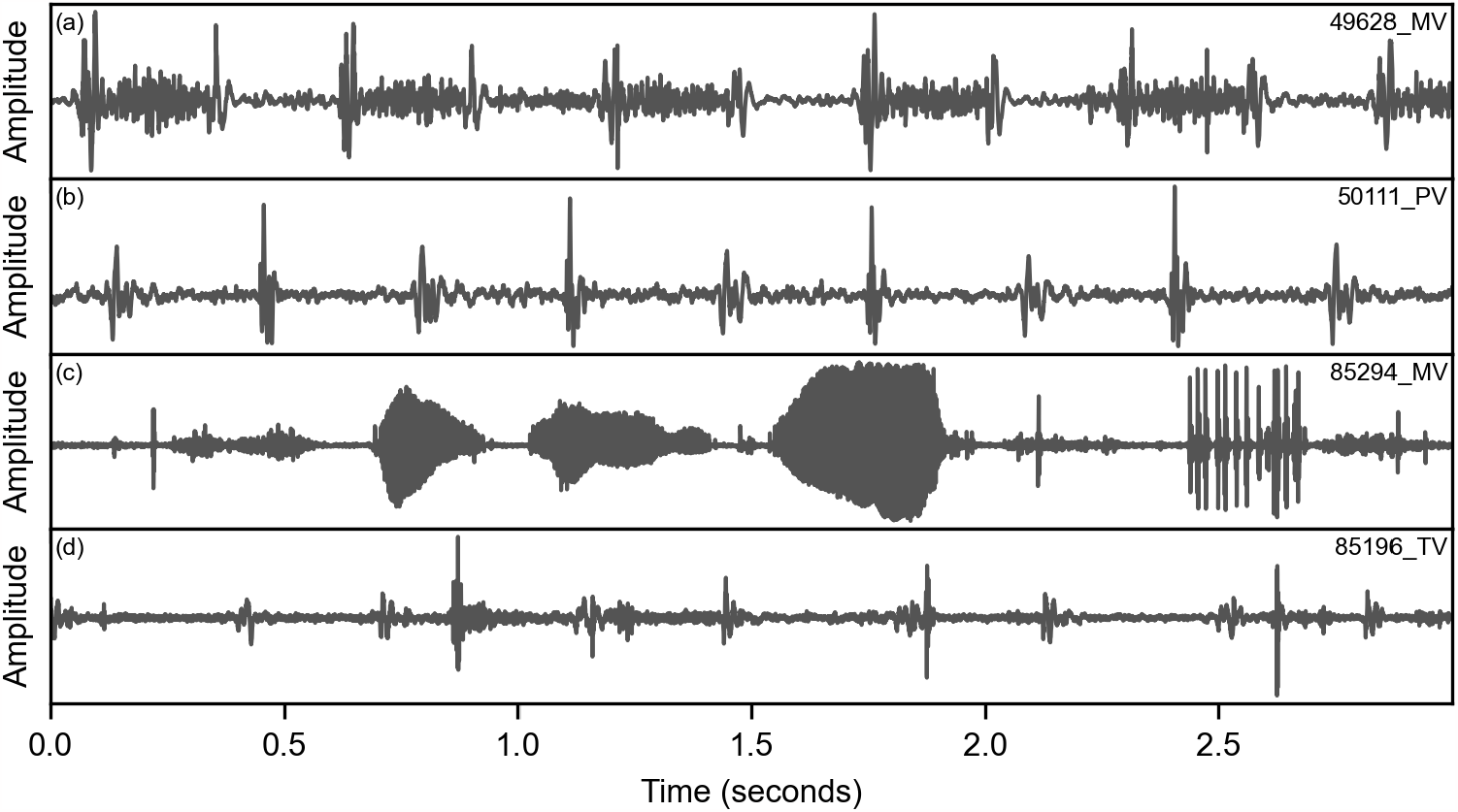
Four example recordings analysed by the algorithm. Three-second windows are shown for each recording, with their database ID shown in the top right. Recording (a) contains a strong systolic murmur that is confidently detected by the algorithm (*C*^(*M*−*N*)^ = 0.25). (b) is a healthy signal that is correctly identified ( *C*^(*M*−*N*)^ = − 0.10). (c) contains significant talking and other noise, and is marked as ‘Unknown’ by the clinician. The algorithm correspondingly rejects the signal with a low confidence of 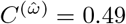. (d) contains a lower amplitude signal with some noise that is marked as ‘Unknown’ by the clinician, but is segmented by the algorithm with a very high confidence of 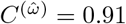.

Results are evaluated via 5-fold cross-validation of the training dataset. Shown are sensitivity (also known as recall), positive predictive value (PPV, also known as precision), and their combined *F*_1_ mean.

### Clinical outcome

A novel part of the 2022 challenge was the use of a custom cost function for the binary clinical outcome task. The challenge organisers argue that traditional metrics for binary classifiers, such as area under the receiver operating characteristic curve, weigh all examples equally and are not optimised for particular clinical contexts [26]. The 2022 cost function was designed to represent the key issues in the deployment of an algorithm in low-cost screening environments [10]. However, one potential limitation of solely using a custom loss function is that results from the challenge cannot be easily compared to other studies applying machine learning to PCG analysis. The cost value can also mask whether the algorithms are actually identifying diagnostic features.

Approximately half of the challenge teams (19) achieved a worse performance on the test set than a random classifier. An optimum random model that achieves a sensitivity of 80% and a specificity of 20% (thus lying on the diagonal line of a receiver operating characteristic, ROC, graph) on the test set would achieve a challenge cost score of 13168. In Fig 7, we compare the training and test scores for all the official challenge entries.

**Fig 7.**
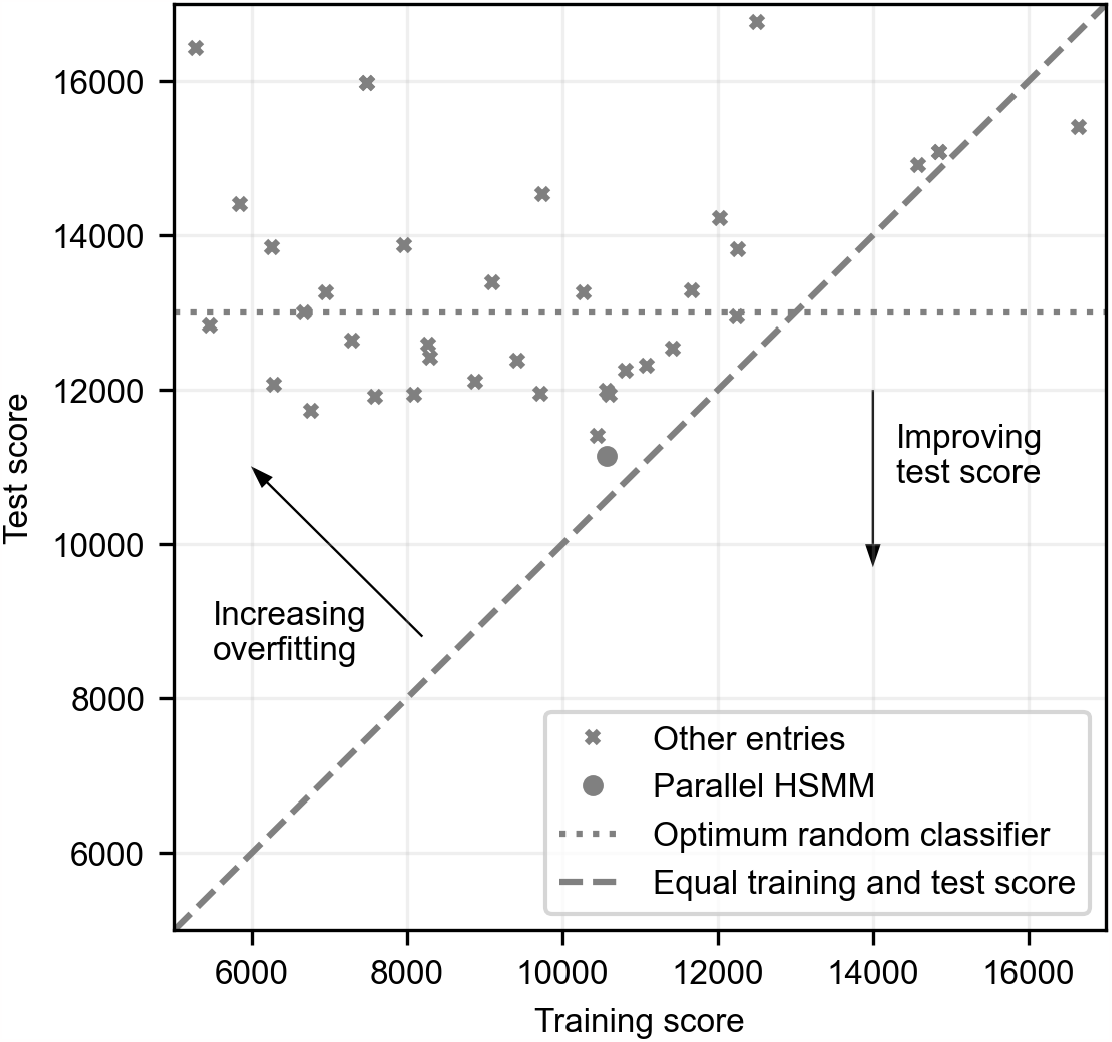
Training and test performance of all the official entries on the clinical outcomes task. A lower score is better. Many algorithms had significantly better performance on the training set, indicating overfitting has taken place. The diagonal line shows ideal performance where training and test performance are equal. The parallel HSMM algorithm achieves the best score and shows little evidence of overfitting. For context, the performance of a random classifier at an optimal operating point is also shown.

Fig 7 also shows that shows that many algorithms overfit to the training set with far worse performance on the final testing set. Many teams used deep learning algorithms commonly deployed in other areas such as speech recognition. However, the small size of the dataset makes training generalisable models a challenge. Although an RNN is used in this work, it is constrained to perform a specific task within the segmentation model and does not generate the final prediction of clinical outcome. Unlike many other teams, our approach does not re-train a completely new model to predict clinical outcome. The CatBoost model was designed to leverage the murmur predictions from the parallel HSMM and combine them with limited patient biometrics. Therefore, it had a limited feature set to train on and the risk of overfitting was low. However, one limitation of this approach is that the murmur detector algorithm is only trained to detect audible abnormal sounds. It is possible that some of the abnormal examples contain inaudible time-frequency features that would be missed by the murmur detection algorithm andhence the CatBoost model. A model trained directly to predict clinical outcome could detect these features.

For a fixed prevalence and dataset size, the cost function can be plotted as a function of sensitivity and specificity as shown in Fig 8. This shows that the overall effect of the cost function is to heavily prioritise sensitivity over specificity. All data on this graph is evaluated on the training set. The graph shows the performance of the clinician murmur ground-truth used to predict abnormal clinical outcome, assuming all ‘Unknown’ cases are referred on as positives. Although the clinician is very specific, the sensitivity is very poor (42%), indicating that many of the patients have heart disease that does not produce an audible signature. The result is a challenge score of 16083. The murmur detection algorithm trades off improved sensitivity for worse specificity but achieves a better score of 13681. However, both of these scores are worse than a random classifier on the random diagonal line of the ROC, which achieves a score of 12579 on the training set.

**Fig 8.**
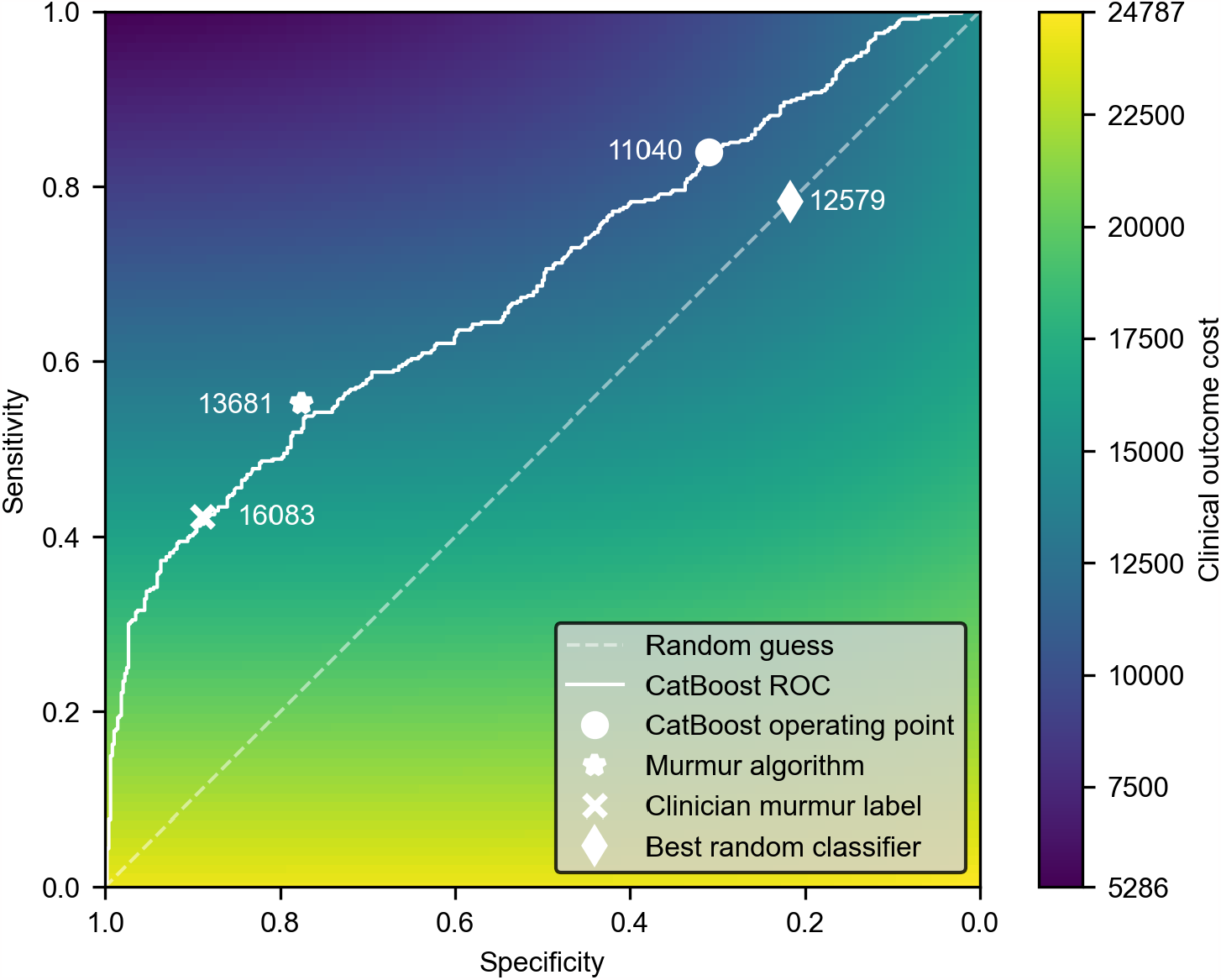
Performance of algorithms and clinician on clinical outcomes task. A receiver operating characteristic (ROC) curve for the CatBoost algorithm at predicting clinical outcome is shown. Also shown are operating points for the clinician (using the murmur label) and the parallel HSMM murmur detection algorithm. The colourmap shows how sensitivity and specificity relate to the PhysioNet challenge cost score. On the training set, an optimum random classifier can achieve a cost score of 12579.

Fig 8 also plots an ROC curve for the performance of our CatBoost algorithm at predicting clinical outcome, with the operating point used in the challenge marked. A score of 11040 is achieved, closely matching the test score of 11144 that won first place. This is an improvement over the optimum random classifier, however, the ROC curve illustrates that the main effect of the CatBoost algorithm has been to shift the operating point from a specific area (i.e. the murmur detection performance) to a sensitive area.

This is highly beneficial to lower the challenge cost function, but may not be practical for widespread screening. Many population-level screening programs prioritise a high specificity over sensitivity [27], because this is crucial to maintaining a high positive predictive value when operating over a low prevalence population. An algorithm with low specificity could lead to a very large number of false positive referrals which would overwhelm secondary care cardiac services, such as echocardiography.

The challenge dataset only provides a binary label of disease and does not provide additional labels on its nature or severity [10]. Given the low sensitivity of clinical auscultation, it is likely that many of the patients recruited have a disease that does not produce audible murmurs or other abnormal sounds. Phonocardiography is a useful tool to detect many structural heart diseases but should be combined with non-invasive cardiac screening tests (e.g. electrocardiography) to provide a more sensitive test for heart dysfunction. It may therefore be beneficial to focus algorithm designs on disease which is known to produce abnormal sounds (e.g. valvular heart disease, septal defects) rather than training models to predict a general abnormality.

## Conclusion

We present a novel algorithm to detect and classify heart murmurs that was the winning entry in the 2022 PhysioNet challenge. The model uses a hybrid approach combining a recurrent neural network with parallel hidden semi-Markov models to accurately segment and classify signals, even in the presence of noise and murmurs. Compared to many other algorithms described in the literature and used in the challenge, our model is lightweight and can easily be interpreted by a clinician.

On the murmur detection task, the model won the first prize in the challenge with a sensitivity of 92.7% and 77.6% for the ‘murmur present’ and ‘murmur absent’ classes respectively. The algorithm also won the first prize in the clinical outcome task. However, its accuracy was reduced compared to the murmur detection task because many of the abnormal patients did not have audible pathological sounds. More specific labels of disease were not available, and future work could investigate the accuracy of these approaches on a per-disease basis.

The algorithm additionally predicts signal quality, so a user can be asked to make a repeat recording if their stethoscope was incorrectly held or if there is substantial bodily or environmental noise. However, the algorithm’s predictions often disagreed with the signal quality label assigned by the clinician. For widespread and low-cost screening, it is essential that heart sound data can be reliably gathered by an unskilled operator.

Future studies should explore the usability of electronic stethoscopes and investigate if automatic signal quality assessment can aid this process.

## Data Availability

All heart sound recordings and clinical data used in this work is available publicly as part of the George B. Moody 2022 PhysioNet challenge (https://moody-challenge.physionet.org/2022/). All code to train and evaluate the algorithm is available in a public repository (https://github.com/am2234/parallel-hsmm-murmur).

## Supporting information

**S1 Fig.**
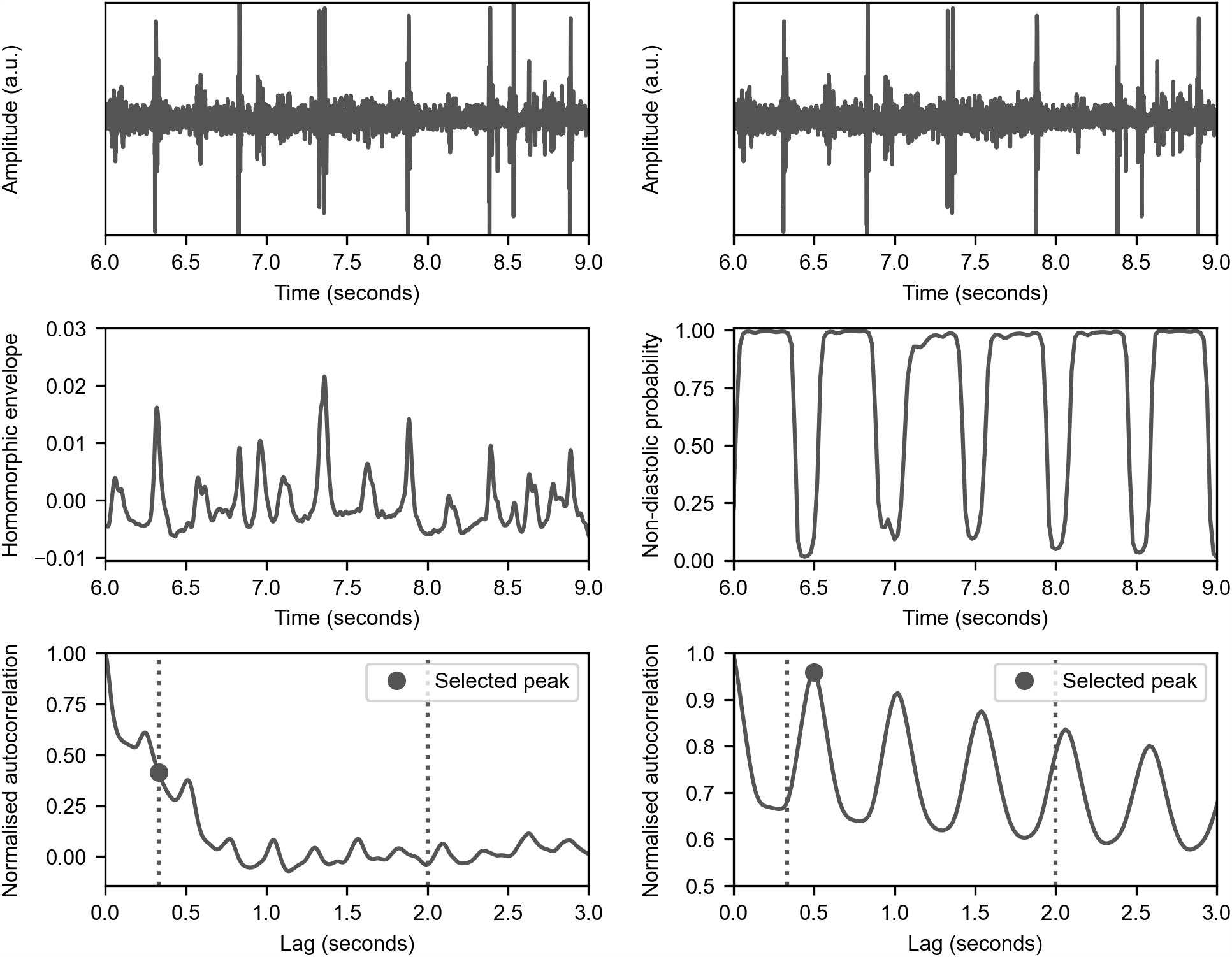
Improved estimate of heart rate using RNN output. Compared here are the methods of Schmidt et al. [24] (left column) and our approach (right column). Schmidt et al. take a homomorphic envelope (middle left) of the signal and then compute its autocorrelation (bottom left). They then search for a peak in a specified range to estimate the heart rate. We use a range of 30-180 bpm for both methods in this dataset because of the faster paediatric sounds. However, this example heart sound (top left) has significant noise which corrupts the envelope and therefore gives a noisy autocorrelation where the correct peak is difficult to find. Our approach instead uses the output of the RNN to create a signal that shows the probability the signal is not in diastole, *P* (*q*_*t*_ ≠ diastole |*x*_1:*T*_, *θ*), (i.e. the summed probability of the S1, S2, systole, and systolic murmur states, middle right). This is a much cleaner signal than the homomorphic envelope, so its autocorrelation (bottom right) is much clearer and the correct peak corresponding to the signal period is easy to find.

## Acknowledgments

Andrew McDonald was supported by the UK Medical Research Council (MR/S036644/1). The first version of the parallel HSMM murmur algorithm was designed by Edmund Kay during his PhD research at Cambridge University Engineering Department. The findings in his thesis inspired many of the improvements in this work.

## Notes

### Competing Interest Statement

AM and AA are inventors on a patent application related to this work (WO2019171021A1).

### Funding Statement

Yes

